# Patient Outcomes and Lessons-Learned from Treating Patients with Severe COVID-19 at a Long-Term Acute Care Hospital

**DOI:** 10.1101/2021.06.10.21255008

**Authors:** Pete Grevelding, Henry C. Hrdlicka, Stephen Holland, Lorraine Cullen, Amanda Meyer, Catherine Connors, Darielle Cooper, Allison Greco

## Abstract

**Objective:** To describe characteristics, clinical management, and patient outcomes during and after acute COVID-19 phase in a long-term acute care hospital in the Northeastern United States.

**Methods:** A single-center retrospective analysis of electronic medical records of patients treated for COVID-19-related impairments, from March 19, 2020 through August 14, 2020, was conducted to evaluate patient outcomes in response to the facility’s holistic treatment approach.

**Results:** 118 admissions were discharged by the data cut-off. Mean patient age was 63 years, 64.1% were male, and 29.9% of patients tested-positive for SARS-CoV-2 infection at admission. The mean (SD) length-of-stay at was 25.5 (13.0) days and there was a positive correlation between patient age and length-of-stay. Of the 51 patients non-ambulatory at admission, 83.3% were ambulatory at discharge. Gait increased 217.4 feet from admission to discharge, a greater increase than the reference cohort of 146.3 feet. 93.8% (15/16) of patients mechanically ventilated at admission were weaned before discharge (mean 11.3 days). 74.7% (56/75) of patients admitted with a restricted diet were discharged on a regular diet.

**Conclusion:** The majority of patients treated at a long-term acute care hospital for severe COVID-19 and related complications improved significantly through coordinated care and rehabilitation.

## INTRODUCTION

Patients hospitalized with severe coronavirus disease-2019 (COVID-19) may face long hospital lengths-of-stay, making it unreasonable to expect a discharge to home without long-term consequences.^1^ COVID-19 is predicted to result in significant morbidity for some patients, with the potential need for medical and rehabilitation services for six-months or longer after the initial diagnosis.^2^ Post-acute care can provide rehabilitation and/or palliative care in the post-COVID phase, as well as provide an alternative to conventional short-term acute care hospitalization (STACH) for active treatment, thereby reducing the burden on the STACH system.^3,4^ The shift of active COVID-19 care from STACHs to long-term acute care hospitals (LTACH) can decrease acute care census during critical stages of the pandemic when resources are limited. It has also been proposed that patients with severe COVID-19 may benefit from inpatient respiratory, functional, and/or neurological rehabilitation.^5^ Early rehabilitation may reduce disability and improve clinical outcomes in patients with COVID-19.^6^ Here, we report patient characteristics, clinical management strategies, and patient outcomes from an LTACH caring for patients with severe COVID-19, as well as make comparisons to the typical medical population cared for at the LTACH.

## METHODS

This retrospective study reports on patients who were treated in regional STACHs for COVID-19 and discharged to an LTACH in the Northeastern United States for post-COVID-19 care and rehabilitation. Study data was collected from March 19, 2020 through August 14, 2020. A reference cohort of 170 patients, who were treated at the same facility during the three months prior (December 1, 2019 through February 29, 2020), was used to compare patient demographics and outcomes (**Table 1**).

**Table 1.** Patient Demographics and Baseline Characteristics.

| Table 1. Patient Demographics and Baseline Characteristics |  |  |
| --- | --- | --- |
|  | COVID-19 Cohort <sup>a</sup> | Reference Cohort <sup>b</sup> |
| Number of individual patients | n = 117 | n = 170 |
| Cohort age, mean (SD), range, years | 63.3 (11.8), 73.0 | 65.6 (14.8), 76.0 |
| Male sex, n (%) | 75 (64.1%) | 98 (57.6%) |
| Male age, mean (SD), range, years | 63.2 (11.4), 64.0 | 63.8 (13.9), 66 |
| Female sex, n (%) | 42 (35.9%) | 72 (42.4%) |
| Female age, mean (SD), range, years | 63.5 (12.7) 66 | 68.0 (15.9), 75 |
| LOS, mean (SD), range, days | 25.6 (13.0), 71 | 29.9 (34.6), 314 |
| BMI, kg/m<sup>2</sup>, mean (SD), range | 30.3 (7.1), 32.9 | 27.4 (7.5), 43.2 |
| Race, n (%) <sup>c</sup> |  |  |
| White/Caucasian | 77 (69.3%) | 143 (87.2%) |
| Non-White/Caucasian: | 35 (30.7%) | 21 (12.8%) |
| Black/African American | 27 (23.7%) | 15 (9.2%) |
| Asian | 7 (6.1%) | 4 (2.4%) |
| Bi/Multiracial | 1 (0.88%) | 2 (1.2%) |
| Two-sided Fisher's Exact test <sup>d</sup> | p=0.0004 |  |
| COVID-19 cohort comorbid conditions at LTACH admission, n (%) <sup>e</sup> |  |  |
| Primary hypertension | 61 (53.0) |  |
| Hyperlipidemia | 49 (42.6) |  |
| Dysphagia | 44 (38.3) |  |
| Type II diabetes mellitus | 41 (35.7) |  |
| Acute kidney failure | 25 (21.7) |  |
| Urinary tract infection | 22 (19.1) |  |
| Severe obesity | 14 (12.2) |  |

LOS: length of stay; BMI: body mass index; SD: standard deviation; CI: confidence interval
<sup>a</sup> Data for the COVID-19 cohort was collected from March 19, 2020 through August 14, 2020.
<sup>b</sup> The reference (typical) population was derived from patients cared for at the facility from December 1, 2019 through February 29, 2020.
<sup>c</sup> Breakdown of self-reported racial demographics. For analysis, the racial demographics were grouped as either White/Caucasian or non-White/Caucasian.
<sup>d</sup> Fisher's Exact test was calculated to determine if the proportion of White/Caucasian and non-White/Caucasian demographics were similarly distributed between the COVID-19 and Reference cohorts.
<sup>e</sup> Comorbid conditions of the COVID-19 cohort were identified by ICD-10 diagnosis codes available in the patient's medical record at discharge from the short-term acute care hospital and admission to long-term acute care.

### Protocols for patients with confirmed or suspected SARS-CoV-2 infection

Similar to arrangements made by other LTACH facilities with regional hospitals, patients who required post-acute care for COVID-19-related issues, and those who were still SARS-CoV-2 positive, were accepted from STACHs to help unburden those facilities.^7^ Additionally, when available beds in the LTACH facility were scarce, healthcare workers and other first responders were prioritized for admission to ensure other regional healthcare facilities were able to be adequately staffed during the pandemic.

Patients with active or prior SARS-CoV-2 infection were housed in separate floors of the hospital, similar to practical arrangements of other post-acute care facilities.^8^ Patients with confirmed or suspected SARS-CoV-2 infection were housed in negative pressure rooms, or in rooms with portable or ceiling-mounted air scrubbers.

Personal protective equipment (PPE) protocols for the COVID-19 cohort included the use of face shields, N95 particulate respirator mask or duck bill surgical mask, scrub caps, boot covers, and uniform laundering at an outside facility; powered air-purifying respirators (PAPRs) were available if needed. Due to a facility shortage of N95 respirator masks (i.e. unknown/unstable resupply chains), these masks were sterilized for reuse by an outside facility.

To decrease personnel exposure to patients with suspected or confirmed SARS-CoV-2 infection and conserve PPE, we developed multidisciplinary “COVID-19 Teams” responsible for patient isolation, testing, implementation of droplet precautions, and cluster care. Further, a dedicated respiratory therapist and intubation box were used to treat patients with active SARS-CoV-2 infection requiring mechanical ventilation or had a tracheostomy.

### Typical care for patients with pulmonary condition

Using standardized measures and functional assessments, interdisciplinary clinical teams evaluated patients to determine functional impairments at admission. When applicable, speech-language pathology (SLP) assessed patients for voicing, swallowing, and cognitive-communication impairments. Patients were mobilized throughout the day, including chair positioning of the bed, transfer to bedside chair, and other exercises/ambulation as appropriate.

Within 24 hours of admission, patients with a tracheostomy were assessed for in-line speaking valve use. As patients progressed with the speaking valve, they were transitioned to tracheostomy capping and placed on the decannulation protocol (**Supplemental Materials**). When appropriate, patients being mechanically ventilated were considered for the ventilator weaning protocol (**Supplemental Materials**); interdisciplinary rounds occur weekly for patients being mechanically ventilated.

#### COVID-19 specific respiratory therapy considerations

SARS-CoV-2 positive patients completed self-directed exercises in their rooms, were seen for individual or co-treatment sessions in room, and, once SARS-CoV-2 negative, participated in group pulmonary exercise therapy and education classes.

Patients who were desaturating or acutely decompensating were treated with proning by a multidisciplinary team including physical therapy, nursing, and respiratory therapy. Prior to implementing proning with patients, staff participated in training sessions on how to safely prone and reposition patients, manage leads and lines, and perform cardiopulmonary resuscitation while in a prone position. Patients who were functionally capable and were previously proned during acute care, were educated on self-proning and encouraged to do so when appropriate.

### Gait/functional status assessment and rehabilitation

At admission, physical therapists evaluated patient ambulatory status by assessing functional ability and gait distance. Patient ‘s received standard individualized physical therapy, and the patient ‘s gait quality and distance was challenged for progression as tolerated. Hypotension and/or tachycardia was present in some patients of the COVID-19 cohort. For these individuals, therapy was aimed at improving tolerance and progression. A modified Functional Independence Measure (FIM) score was used to describe functional ability as the amount of assistance provided by the therapist(s) during treatment (**Supplemental Table 1**).

### Speech-language pathology

Many patients in the COVID-19 cohort presented with cognitive-communication deficits, potentially as a result of COVID-19 induced hypoxia, prolonged intubation, and/or sedation.^9^ When appropriate, cognitive-communication assessments were performed by an SLP on the COVID-19 team. Using tools such as the Bioness Integrated Therapy System (BITS®), worksheets, and group therapy sessions, SLP sessions focused on attention, memory, functional skills, and compensatory strategy use. A modified National Outcomes Measure System (NOMS) assessment was used to summarize the patient ‘s overall cognitive communication status at admission and discharge (**Table 3**).

Due to the correlation between prolonged intubation and dysphagia, SLP interventions also targeted swallowing dysfunction.^10^ Dysphagia management comprises several aerosol-generating procedures, including oral mechanism examination, cough testing, reflexive cough, swallowing trials, and secretion management. Given the proximity and prolonged exposure to aerosols during instrumental evaluations, and the need for multiple staff members, procedures such as Fiberoptic Endoscopic Evaluation of Swallow or Modified Barium Swallow Study were minimized. Thus, SLPs heavily relied on clinical swallowing evaluations for patients with active SARS-CoV-2 infection.Additionally, some patients in the COVID-19 cohort consented to performing clinical swallowing evaluations via telehealth to reduce potential SARS-CoV-2 exposure and transmission.

### Statistical analysis

Data was analyzed using GraphPad Prism version 9.0.0 (GraphPad Software, San Diego, CA). Prior to analysis, data was tested for normality using the Shapiro-Wilk test and QQ-plot visualization; one or more group from each datasets was abnormally distributed (p<0.05), so nonparametric testing was used during analysis.

For hypothesis testing between two unpaired groups, Mann-Whitney rank comparison test was conducted; for paired two group testing, Wilcoxon matched-pairs signed rank test was conducted. For hypothesis testing between three groups, Kruskal-Wallis analysis of variation test and Dunn ‘s multiple comparison test were conducted. Fisher ‘s exact test was calculated to compare the observed proportions of patients within the COVID-19 cohort, which self-reported as white versus non-white, to the Reference Cohort.

Nonlinear regression (NLR) was conducted to determine correlation between two conditions using least-squares regression; 95% CIs are reported. Extra sum-of-squares F-test was completed to evaluate the calculated slope of each NLR against a hypothetical slope=0. This descriptive retrospective study should be assessed while keeping study limitations in mind, including, adoption and adaptation of procedures throughout the study period, and the use of medical records to collate data. This study is strengthened by the breadth of quantitative outcomes and the detailed descriptions of potential presentations and complications that can be expected for patients with COVID-19 being treated in a LTACH.

## RESULTS

### Patient demographics

During the study period, 117 individuals were admitted for COVID-19 or post-COVID-19 related care. COVID-19 admissions peaked during May 2020 (**Figure 1A**), approximately four-weeks later than the New England/New York City area.^11,12^ Due to acute decompensation requiring multiple, temporary STACH readmission(s) followed by LTACH readmission, eight individual patients accounted for ten additional admissions (10/127, 7.9%). Of the 127 total admissions, 118 were discharged by the data cut-off, with 9 still receiving care. The COVID-19 cohort represented 17.2% (127/737) of the hospital census during the 4.5-month/148-day study period. For each admission, the mean (SD, range) STACH length of stay (LOS) prior to LTACH admission was 35.5 (22.6, 4-122) days. The mean LOS at LTACH was 25.5 (13.0, 1-72) days. NLR analysis indicated that there was no correlation between STACH LOS and LTACH LOS (**Figure 1B**). Further, the mean LOS of the COVID-19 cohort was similar to the reference cohort LOS of 29.9 (34.6, 1-315) days (**Figure 1C**).

**Figure 1.**
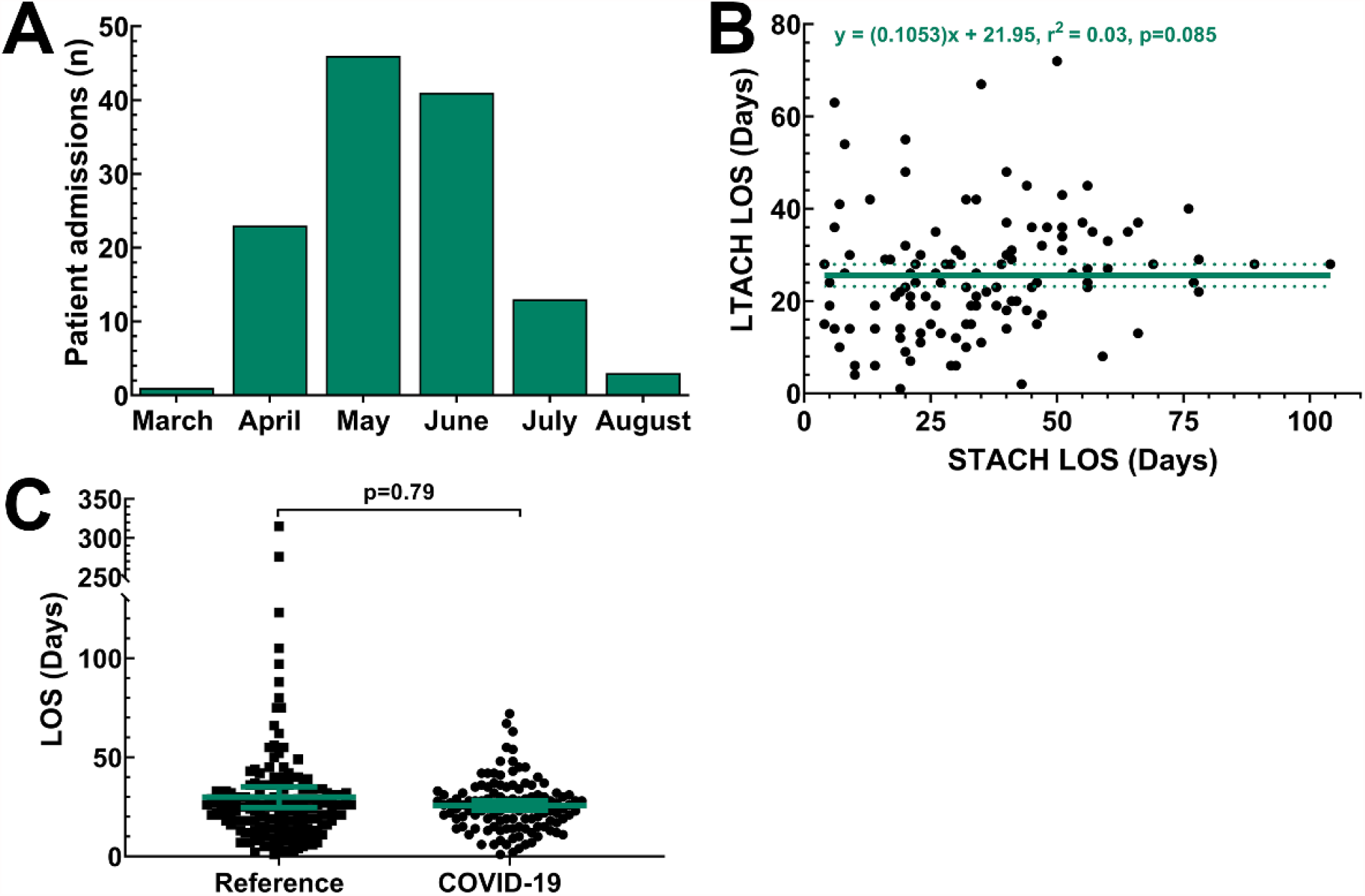
Trends in patient admission and length of stay (LOS) during the COVID-19 pandemic. **(A)** From March 19, 2020 through August 14, 2020, 117 patients were admitted to the long-term acute care hospital (LTACH) for the treatment and rehabilitation of COVID-19 illness related conditions. 10 patients were readmitted to the LTACH after a temporary-transfer back to short-term acute care, resulting in a total of 127 admission during the study period. The highest frequency of admissions occurred in May 2020. **(B)** Nonlinear regression showed no correlation between patient LTACH LOS and STACH LOS. Solid regression line shows the correlation coefficient, and the dotted lines show the 95% confidence interval (CI). **(C)** The mean LTACH LOS was similar between the COVID-19 cohort and the reference cohort treated at the same facility during the prior 3 months. **C**, the lighter colored lines represent the mean and the 95% CI.

Compared to the reference cohort (170 individual patients), the COVID-19 cohort (117 individual patients) had similar proportions of males-to-females, was younger (p=0.30, **Figure 2**), and had a greater representation of non-white racial demographics (Black/African American, Asian, Bi-/Multiracial) (p=0.0004, **Table 1**). Further, the COVID-19 cohort had a significantly higher body mass index (BMI) than the reference cohort (p=0.0005, **Table 1** and **Figure 2**). The most common comorbidities present at admission were hypertension (53.0%), hyperlipidemia (42.6%), and dysphagia (38.3%) (**Table 1**).

**Figure 2.**
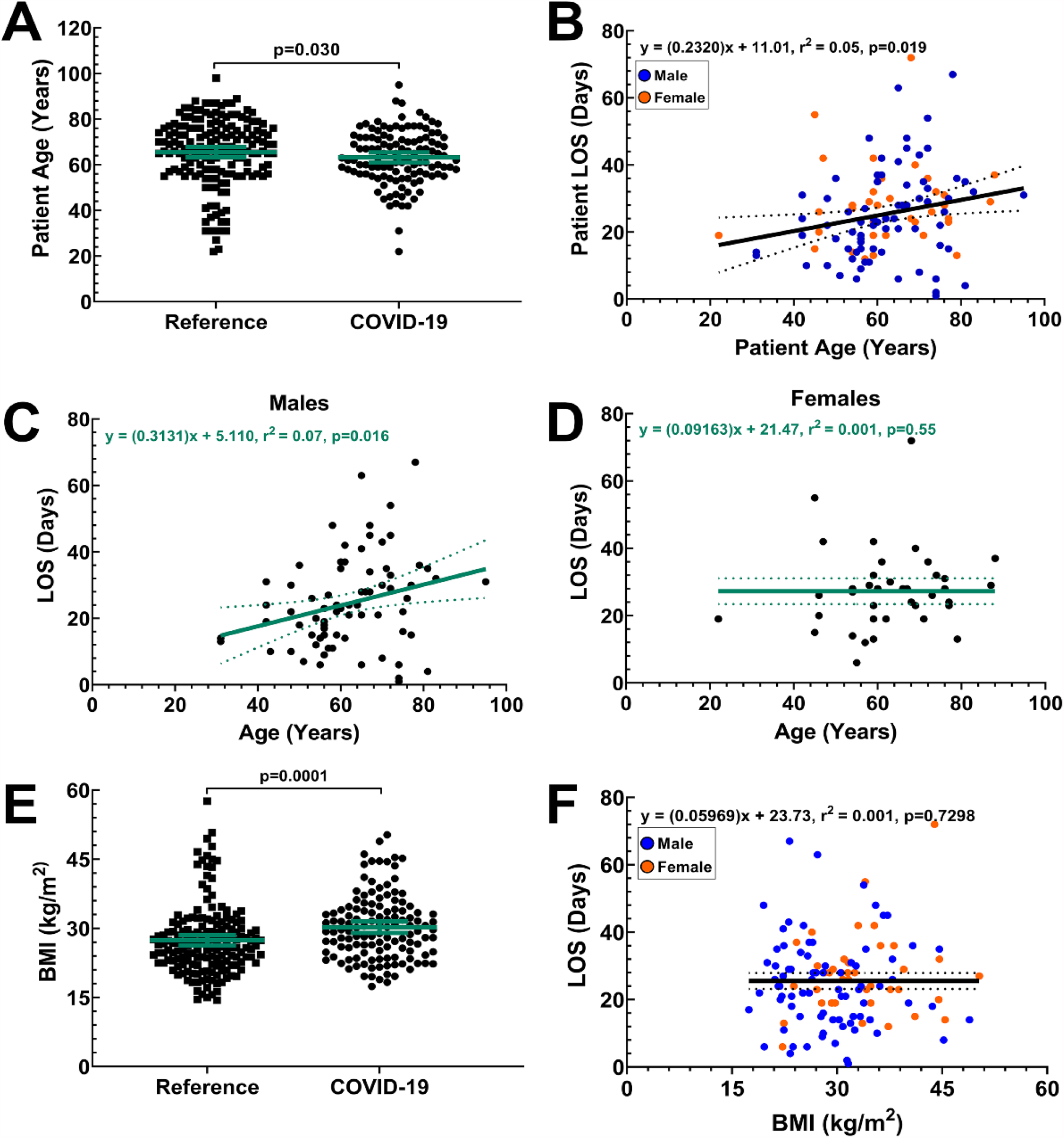
Age and BMI as risk factors for prolonged COVID-19 illness. Shown are data for 127 COVID-19 cohort admissions that were discharged by the data cut off; this includes data for patients who were admitted and discharged multiple times throughout the study. **(A)** Mean age of the COVID-19 cohort and Reference cohort were significantly different. **(B-D)** Nonlinear regression identified a positive correlation between patient age and LTACH LOS in the overall COVID-19 cohort **(B)**, a pattern also observed in males only **(C)**, but which was not present in females only **(D)**. Reference and COVID-19 cohorts BMIs were significantly different at admission **(E)**. Non-linear regression showed no correlation between patient BMI and LTACH LOS **(F). A and E**, lighter colored lines represent the mean and 95% CI. **B, C, D, and F**, solid regression lines show the correlation coefficient, surrounded by the 95% CI in dotted lines.

### Outcomes

The mean age of the COVID-19 cohort was younger than the reference cohort (p=0.030, **Figure 2A**). Using LOS as a read-out for disease severity (i.e., the more severe the COVID-19 illness, the longer the length of stay in rehabilitation), NLR analyses was performed to determine if patient sex, age, or BMI affected LOS, all of which have been noted to increase the risk of severe or prolonged COVID-19 illness.^13–15^ Examining age as a potential risk factor for longer LOS and prolonged COVID-19 rehabilitation, we observed a positive correlation among COVID-19 patients (**Figure 2B**). When each sex was analyzed separately, we observed a positive correlation in males of the COVID-19 cohort (**Figure 2C**), but not females (**Figure 2D**). Despite the mean BMI of the COVID-19 cohort being higher than the reference cohort (**Table 1, Figure 2E**), we observed no correlation between COVID-19 cohort patient BMI and LOS overall (**Figure 2F**), nor for males or females separately (data not shown).

#### Respiratory therapy

Of the 43 patients admitted with a tracheostomy, 37.2% (16/43) required mechanical ventilation and 62.8% (27/43) did not; 93.8% (15/16) of mechanically ventilated patients in the COVID-19 cohort were weaned by the data cut-off. Compared to the reference cohort, the mean (SD) ventilator wean time of the COVID-19 cohort tended to be shorter than the reference [21.6 (11.1) versus 11.3 (8.4) days; p=0.23]. Given the small number of patients being mechanically ventilated in the reference cohort (n=7), we also compared the COVID-19 cohort wean time to all patients for fiscal year 2019 (FY19) in the LTACH [12.2 (9.8) days; n=37] and found no difference between the two (p>0.99, **Figure 3A**). For those weaned from mechanical ventilation, it was an additional 15.1 (13.3) days until tracheostomy decannulation. In comparison, for those not being mechanically ventilated, the mean time from admission to tracheostomy decannulation was 16.3 (11.4) days

**Figure 3.**
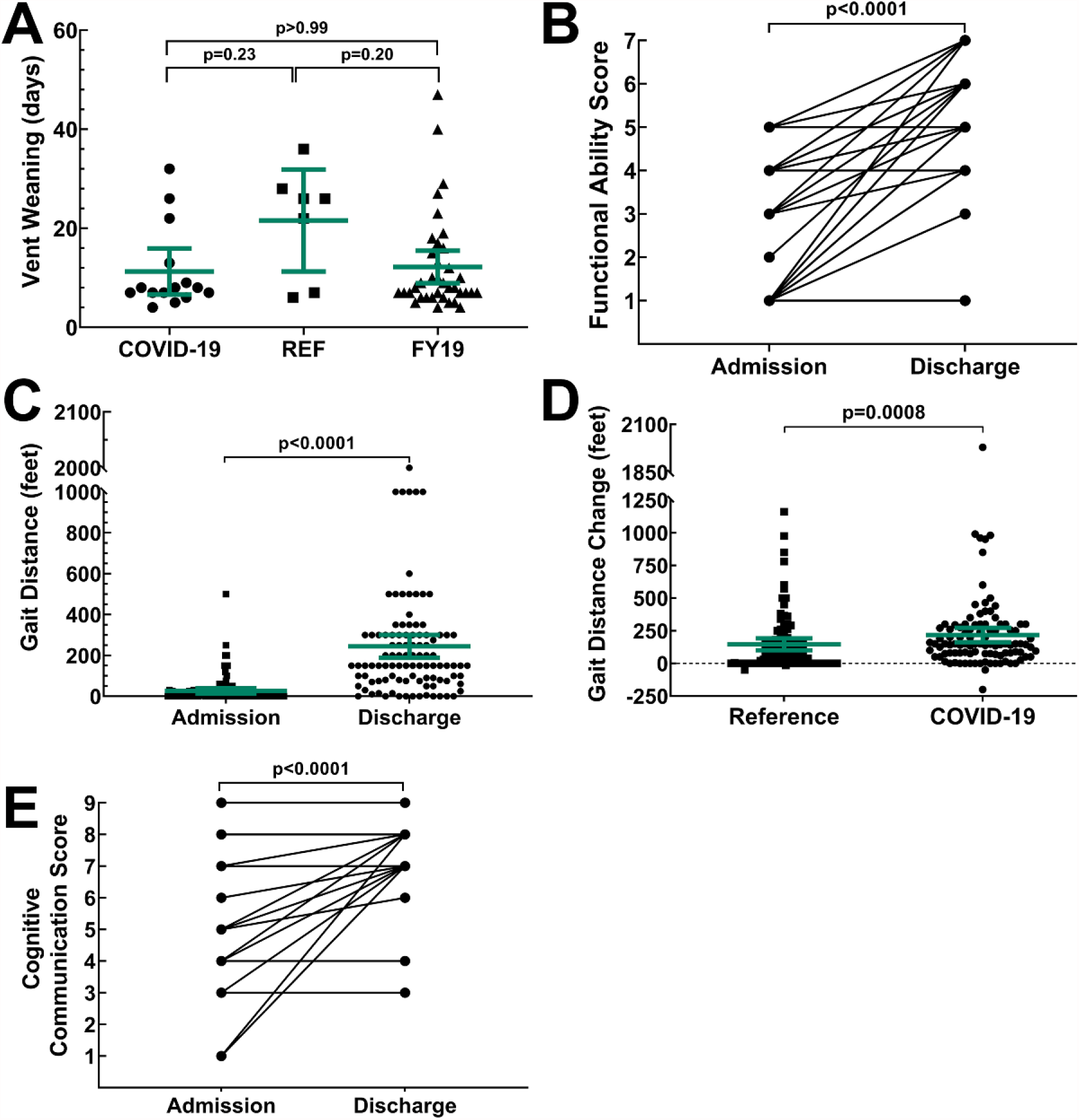
Patient respiratory, functional ability, and cognitive-communication outcomes. **(A)** Ventilator wean time was not significantly different between the COVID-19 cohort, reference cohort, and all patients being mechanically ventilated during fiscal year 2019 (FY19). At both admission and discharge, patients were evaluated on their functional ability **(B)** and the distance they were able to ambulate **(C)**. The change in gait distance from admission to discharge was compared between the Reference and COVID-19 cohort to determine if the extent of improvement between the two groups was comparable **(D)**. At admission and discharge, the cognitive communication score of patients recommended for SLP services was also evaluated **(E). A, C, and D**, lighter colored lines represent the mean and 95% CI.

#### Physical and occupational therapy

Due to wheelchair dependence prior to STACH admission, patient readmission to STACH, continuing-care at the time of data cut-off, and/or incomplete data collection, complete gait and functional status data was available for 99/127 total admissions. At LTACH admission, using the modified FIMs score (**Table 2**), 51 patients were unable to ambulate, 1 needed maximal assistance, 6 needed moderate assistance, 35 needed minimal assistance, and 6 needed supervision. The vast majority (86.9%; 86/99) displayed functional status improvement from admission to discharge, with many patients increasing by 4 or more functional ability levels (p<0.0001, **Figure 3B**). From admission to discharge, the mean (SD) increase in ambulating distance was 217.4 (279.6) feet (p<0.0001, **Figure 3C**); this was a greater improvement than the reference cohort at 146.3 (221.6) feet (p=0.0008, **Figure 3D**). Of the 51 patients who were non-ambulatory at admission, 40 (83.3%) patients were ambulatory by LTACH discharge. Only 11 patients remained unable to ambulate at discharge, 3 of whom were acutely readmitted to a STACH facility.

**Table 2.** Functional Ability Score^a^.

| Table 2. Functional Ability Score<sup>a</sup> |  |  |  |
| --- | --- | --- | --- |
| Description | Score | Admission (n) | Discharge (n) |
| **Unable/Dependent:** patient is either unable to ambulate or is only able to assist up to 24% of activity. | 1 | 51 | 11 |
| **Maximal Assistance:** patient is able to assist with 25% - 49% of activity. | 2 | 1 |  |
| **Moderate Assistance:** patient is able to assist with 50% - 74% of activity. | 3 | 6 | 3 |
| **Minimal Assistance:** patient performs at least 75% of activity. | 4 | 35 | 14 |
| **Supervision:** patient does not need physical assistance but does require hands-on guidance, supervision for safety, cueing, coaxing, or set up. | 5 | 6 | 29 |
| **Modified Independence:** does not need the physical presence of a second person, however, requires equipment, takes more than reasonable time, or there are safety concern. | 6 | 0 | 28 |
| **Independence:** does not require any equipment or the physical presence of a second person. | 7 | 0 | 14 |
| Patients (n) |  | 99 | 99 |
| Mean Score (±SD) |  | 2.4 (±1.5) | 4.9(±1.7) |
| p-value<sup>b</sup> |  | <0.0001 |  |
| <sup>a</sup>To track patient functional ability, we used a modified FIM score to assess the functional ability and level of assistance required at patient admission and discharge. |  |  |  |
| <sup>b</sup>Calculated using the paired Wilcoxon matched-pairs signed rank test. |  |  |  |

#### Speech-language pathology

59.1% (75/127) of the COVID-19 cohort admissions were recommended for SLP evaluation. Of those, 81.3% (61/75) were admitted with a modified diet or instructions for nothing by mouth (NPO). Following a dysphagia evaluation, most patients were upgraded from NPO to a regular consistency diet. At discharge, 74.7% (56/75) of patients were consuming a regular consistency diet. Further, 49.3% (37/75) of patients evaluated by SLP were admitted with a tracheostomy, with or without mechanical ventilation, and 73.0% (27/37) were found to have some form of voicing disorder, including aphonia (13/37), dysphonia (13/37), or dysarthria (1/37). At discharge, only 35.1% (13/37) of patients had voicing limitations.

SLPs also evaluated patients for cognitive-communication deficits using the modified NOMS scale shown in **Table 3**. At admission, 58.7% (44/75) of patients were rated as either baseline or within functional limits, 37.3% (28/75) were found to have impairments ranging from mild to severe, and 4% (3/75) were unable to be assessed; the mean (SD) cognitive-communication score at admission was 7.2 (1.9). Deficits primarily affected the areas of attention, processing speed, short-term memory, and complex executive functioning skills. Many patients showed improvement by discharge with 72% (54/75) being at baseline or within-functional-limits, 21.3% (16/75) had only mild residual cognitive deficits needing minimal cues or memory aides for maintaining attention, completing tasks, or problem solving; 6.7% (5/75) of patients were discharged with continuing moderate-to-severe cognitive deficits. At discharge, the mean (SD) cognitive-communication score was 7.8 (1.1), a modest, yet significant, improvement from admission (p<0.0001, **Figure 3E**). Continued SLP services were recommended for 39% of patients after discharge.

**Table 3.**
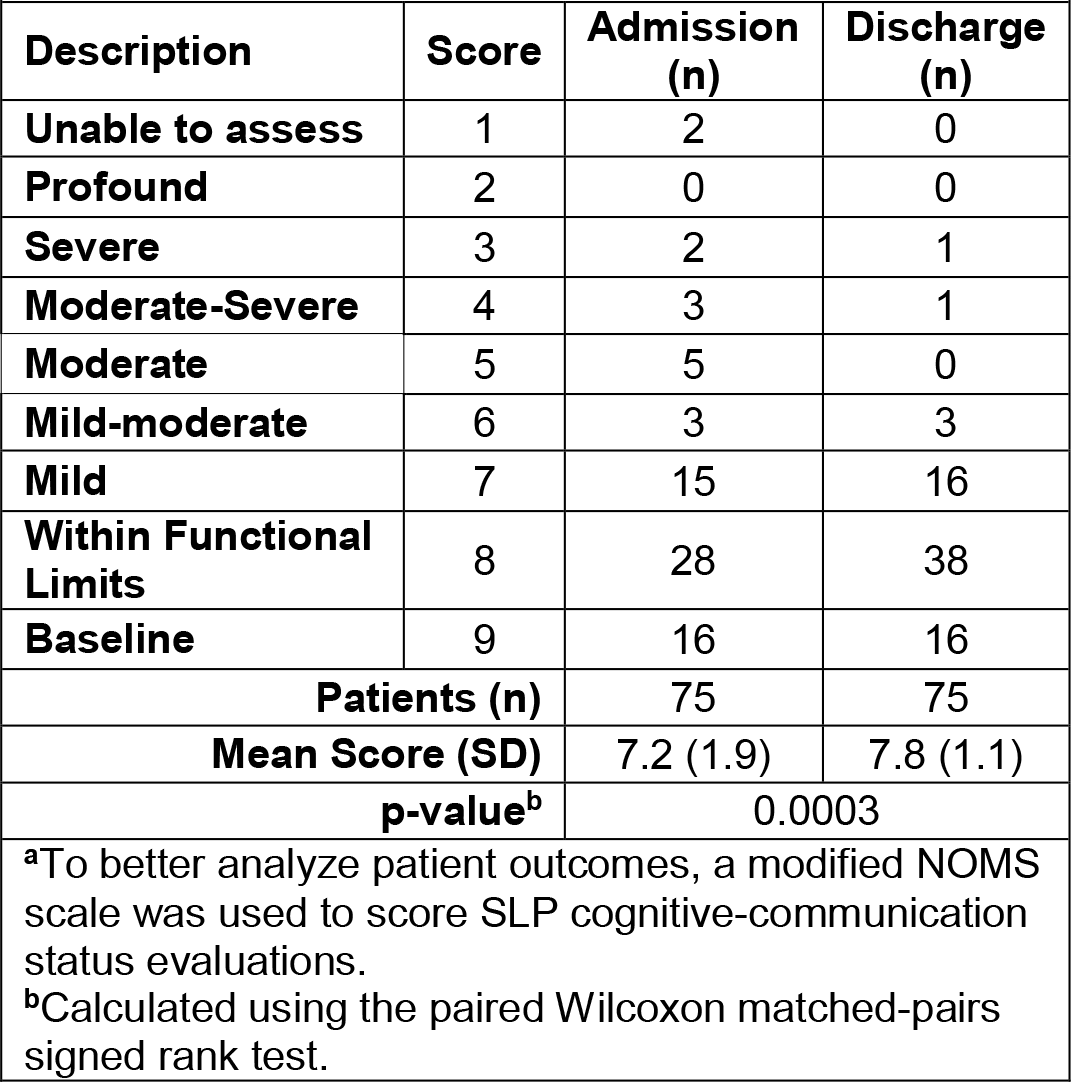
Cognitive-Communication Status Score^a^.

| Table 3. Cognitive-Communication Status Score<sup>a</sup> |  |  |  |
| --- | --- | --- | --- |
| Description | Score | Admission (n) | Discharge (n) |
| Unable to assess | 1 | 2 | 0 |
| Profound | 2 | 0 | 0 |
| Severe | 3 | 2 | 1 |
| Moderate-Severe | 4 | 3 | 1 |
| Moderate | 5 | 5 | 0 |
| Mild-moderate | 6 | 3 | 3 |
| Mild | 7 | 15 | 16 |
| Within Functional Limits | 8 | 28 | 38 |
| Baseline | 9 | 16 | 16 |
| Patients (n) |  | 75 | 75 |
| Mean Score (SD) |  | 7.2 (1.9) | 7.8 (1.1) |
| p-value<sup>b</sup> |  | 0.0003 |  |
| <sup>a</sup>To better analyze patient outcomes, a modified NOMS scale was used to score SLP cognitive-communication status evaluations. |  |  |  |
| <sup>b</sup>Calculated using the paired Wilcoxon matched-pairs signed rank test. |  |  |  |

#### Additional wound care, physical therapy, and medical service considerations

With proning becoming a standard of care for COVID-19-related respiratory failure and pneumonia, many patients in the COVID-19 cohort developed atypical facial pressure injuries during their STACH stay. Patients in the COVID-19 cohort were admitted with approximately 69 total body pressure injuries (Stage 3 or 4) requiring consultation; 30% were located on the face, usually on both cheeks, with one more severe than the other and having thick eschar development. Conservative treatment without sharp debridement resolved most cases of facial pressure injuries. New injuries were prevented by implementing adhesive foam cushioning to facial pressure areas. Patients were also likely more hemodynamically stable during LTACH care and therefore somewhat less likely to develop pressure injuries.

Unilateral and bilateral wrist and foot drop was also observed in some patients, potentially due to prolonged prone positioning in the STACH causing peripheral nerve compression. Patients with wrist drop made some improvement, though some required orthoses or occupational therapy after discharge. Some COVID-19 patients presented conditions atypical to respiratory disease, such as neurological findings, peripheral nerve injuries, paresthesia, and/or cognitive impairment. Neurological symptoms may have been a result of the use of paralytics or prolonged prone positioning during STACH treatment.^9,16^

## DISCUSSION

Emergence of novel SARS-CoV-2 and the resultant COVID-19 disease has resulted in a worldwide pandemic with millions of infections and nearly 1.5 million deaths as of this writing.^17^ For other facilities to reference now or in the future when treating patients with COVID-19, the goal of this retrospective study was to summarize and report the observations, experiences, and methods used by clinicians at our LTACH and how these practices impacted patient outcomes. Using our holistic treatment strategy, we focused on all aspects of patient recovery, with the majority of our patients with severe active-COVID-19 or post-COVID-19, showing significant improvement through this coordinated care.

During the study period, 93.8% of patients admitted on mechanical ventilation were weaned, and 96.3% of patients admitted with a tracheostomy without mechanical ventilation were decannulated. Though many patients had functional limitations and were non-ambulatory at admission, the COVID-19 cohort showed significant functional improvement by discharge, including a 149% greater change in gait-distance travelled compared to the reference cohort. Patients receiving speech-language therapy also showed improvements during their LTACH treatment, with 40.5% fewer patients having voicing limitations at discharge and only 28% having residual cognitive-communication deficits. Together, these observations indicate the potential benefit of individualized, focused, and holistic rehabilitation in a population severely affected by COVID-19.^18^

Though not significant, the COVID-19 cohort ventilator wean time (10.4 days) was shorter than historical facility wean times (12.2 days in 2019, 20.6 days in 2018, and 14 days in 2017).^19^ Based on our clinical observations, the COVID-19 cohort generally presented fewer complicated pulmonary and cardiac comorbidities than typical patients with tracheostomy, with or without mechanical ventilation. This may have contributed to the shorter ventilator wean time. These observations support the idea that pulmonary rehabilitation could play an important role in COVID-19 treatment and recovery.^20^ Further, compared to patients with chronic pulmonary conditions, the COVID-19 cohort patients, who were generally new to respiratory deficits, improved rapidly with appropriate respiratory management.

In regards to patient susceptibility and risk for severe COVID-19 illness, we observed a positive correlation between patient age and patient LOS. In contrast to what has been reported, we did not observe a correlation between patient BMI and disease severity/LOS.^13–15^ These differences could be attributed to several reasons, including better pre-COVID-19 health status compared to patients typically cared for at the facility, current employment status at the time of COVID-19 diagnosis (many of the cohort were healthcare workers or first responders), or motivation to return home (as visitation was restricted).

The quick progression in cognitive-communication skills during LTACH stay was also likely multifactorial, involving discontinuation of sedatives, improved metabolic status, awareness of deficits, and an ability for patients to carry over compensatory strategies learned in therapy. However, ongoing cognitive-communication impairment is possible in patients who have had COVID-19, and these individuals may benefit from continued therapy services after discharge.^21^

The suspected reason many of our patients were admitted on a modified diet or NPO was due to either an inability for the patient to participate in swallowing assessments at acute care, the severity of their medical condition, or the limited access to instrumental assessments during SLP evaluations due to droplet precautions. The prompt advancement of diet in the LTACH setting was mostly the result of clinical swallowing evaluations showing minimal residual weakness within the oropharyngeal swallowing mechanism. Therefore, it may be possible to largely rely on clinical swallowing evaluations for patients with COVID-19, thus minimizing the risk of viral exposure by limiting aerosol-generating instrumental assessments.^18^ To protect from aerosols when assessing patients with unknown or suspected positive SARS-CoV-2 status, SLPs should consider the continued use of clear face masks, face shields, and/or other eye protection during therapy sessions.

Patients also likely received emotional benefit from the formation of inpatient COVID-19 support groups. These groups, facilitated by a physical therapist and a social worker, were a collaborative effort to provide patients who were recovering from COVID-19 with the opportunity to speak with other patients experiencing similar concerns during their hospital course. With guidance from the group facilitators, patients were encouraged to ask questions and share their experiences in an open discussion format, which ultimately generated insightful feedback to the staff on patient care during the pandemic. Conversation topics focused on: processing the initial illness onset and acute hospital stay; acknowledging and learning to cope with their physical, respiratory, emotional, and social changes; and preparing for their future after LTACH discharge. Participation was capped at 6 patients per meeting and multiple meetings were convened as necessary to accommodate all interested patients.

### Mitigating SARS-CoV-2 transmission in “non-COVID-19 patient population”

Patients cared for at LTACHs typically have complex medical conditions and are at increased risk for infection and fever, thus there was a pressing need to isolate any potential source of SARS-CoV-2. Despite what has been described as “typical” COVID-19 symptoms, patients presented with a spectrum of respiratory symptoms, from asymptomatic to respiratory distress. Consequently, all febrile patients were required to undergo SARS-CoV-2 testing and were isolated with droplet precautions until ruled-out. With only one exception, all non-COVID-19 patients tested negative for SARS-CoV-2, indicating our protocols effectively isolated the 37 patients admitted with active SARS-CoV-2 infections. Our observation supports preemptive testing in LTACH and other healthcare facilities to lower the incidence of SARS-CoV-2 transmission.^22^ Given the documented issues of SARS-CoV-2 transmission in some long-term care facilities, it is possible to imagine what the alternative may have been without preemptive testing.^23–25^

One limiting aspect of care during this period of the pandemic was the length of time it took to obtain SARS-CoV-2 test results for patients who were admitted with an active infection, so they could come off droplet precautions, which in many cases was over two months.^26^ On May 20, 2020, Connecticut Department of Public Health released a memo supporting their agreement with the Centers for Disease Control and Prevention findings that live virus was undetectable after 9 days of infection, allowing for the use of a symptom-based-strategy rather than the test-based-strategy.^27,28^ We implemented a more conservative approach, requiring at least 14 days since diagnosis and 5 days without fever or evolving symptoms. Further, given the low facility infection rate of the non-COVID-19 population, the facility policy changed around the same time from transferring patients under investigation to the COVID-19 floor, to ruling-out in place with the use of droplet precautions and a portable room air-scrubber. Coming off droplet precautions was instrumental in getting patients out of their rooms and having full access to therapy.

It was also evident early-on that regular, clear, and transparent communication was, and still is, vital for staff acceptance of the constantly changing situation, guidelines, and personal protective equipment protocols. To support this, department directors and managers devoted time each day to discussing COVID-19–related patient issues. These directors then met weekly with key staff members to further discuss the issues and disseminate information. Further, emails were frequently sent to all employees detailing COVID-19–related changes, statistics, and other topics of interest. In person communication was also helpful in correcting rumors and serving as a forum for establishing best practices in the ever-changing situation.

In conclusion, to alleviate crowded and overwhelmed STACH facilities, we envision the strategic use of LTACHs earlier in a patient ‘s hospital course to treat and rehabilitate those with severe COVID-19. With a greater understanding of rehabilitation progression, clinical care can be adapted to maximize the recovery of this population.

## Data Availability

Requests for copies of de-identified study data will considered on a case by case basis.

## DECLARATIONS

### Ethics Approval

This study was written in compliance with our institutional HIPAA policy and the standards set by the Declaration of Helsinki. Prior to beginning, this retrospective study was reviewed and given exempted status by the Gaylord Specialty Healthcare Institutional Review Board.

### Conflicts of Interest

Authors have no conflicts of interest to declare.

## Acknowledgements

Medical writing and editorial assistance was provided by Agnella Izzo Matic, PhD, CMPP (AIM Biomedical, LLC) and funded by Gaylord Specialty Healthcare. The authors want to acknowledge the dedication, compassion and expertise of our staff during this pandemic and, as always, we want to acknowledge our patients who inspire us to be better every day.

## Abbreviations

BMI: Body mass index
COVID-19: Coronavirus disease-2019
LOS: Length of stay
LTACH: Long-term acute care hospital
NPO: Nothing by mouth
SARS-CoV-2: Severe acute respiratory syndrome coronavirus 2
SLP: Speech language pathology
STACH: Short-term acute care hospital

## Notes

### Competing Interest Statement

The authors have declared no competing interest.

### Funding Statement

No external funding was received to complete this study.

### Author Declarations

Prior to beginning, this retrospective study was reviewed and given exempted status by the Gaylord Specialty Healthcare Institutional Review Board.

