## Supplemental Materials for "Patient Outcomes and Lessons-Learned from Treating Patients with Severe COVID-19 at a Long-Term Acute Care Hospital"

### SUPPLEMENTARY MATERIALS

#### Mechanical ventilation weaning protocol

To be a candidate for Spontaneous Breathing Trial, the patient must pass the following criteria:

1. **Clinical Assessment prior to Spontaneous Breathing Trial (SBT):**
    - **a.** Successful Spontaneous Awakening Trial (respiratory therapy [RT] to confirm with nursing [RN])
    - **b.** No hemodynamic instability (RT to confirm with RN)
    - **c.** Assess mental status to be within range of alert, calm, mild drowsiness to mild restlessness
    - **d.** $PaO_2/FiO_2$ greater than 200 (Obtain values from most current ABG- done within last 48 hours)
    - **e.** PEEP $\leq$ 7.5
    - **f.** $FIO_2 \leq 0.50$
    - **g.** Patient is making efforts to breathe
    - **h.** Secretions scant non-purulent, suctioning not more than every 4 hr
    - **i.** Temp $\leq$ 100.5°F
    - **j.** Document tracheostomy size and type

**Prior to initiation of SBT, notify cardiac monitor technician**

2. **Interventions:**
    - **a.** If patient had completed 2 hrs or more of trach mask trial at acute care, can proceed with supervised 30-minute tracheostomy mask trial. If patient remains stable, proceed with tracheostomy mask progress.
    - **b.** If patient has not performed tracheostomy mask trial, place on CPAP of +5 and PSV of +5 with a maximum $FIO_2$ of 0.50 and perform RSBI (Rapid Shallow Breathing Index)
        - Respiratory rate/tidal volume (f/Vt) should be <105 to proceed with tracheostomy mask trial
    - **c.** If patient is on CPAP and remains stable, continue on current settings: PS 5, CPAP 5, $FIO_2 < 0.50$. Then if these settings are tolerated for 30 minutes to 2 hours (based on overall status of patient), initiate reassessment for spontaneous breathing success. If patient continues to remain stable, begin with tracheostomy mask trials.
    - **d.** Resume previous ventilator support settings & terminate spontaneous breathing trial and inform RN/MD, if any of the following occur: Respiratory Rate > 35; Respiratory Rate <8; Oxygen saturation <88%; Respiratory distress; Mental status change; Acute cardiac arrhythmia/Hemodynamic changes; Temperature ≥ 100.5°F
    - **e.** Obtain ABG per ABG Protocol.

3. **Reassessment:**

Spontaneous breathing trial deemed successful based upon the following criteria: Stable gas exchange; Hemodynamically stable; Temperature ≤ 100.5 °F; RR < 35; SpO2 > 88% on not more than 50% $FiO_2$; Positive cuff leak; Absence of diaphoresis and absence of use of accessory muscles; Ability to maintain airway with ability to clear secretions

**If patient fails any criteria in reassessment, pulmonary physician will be called, and trial will be attempted next day.**

- Assess for presence of cuff leak
- Begin 2 hr TM trial BID with intervening 2 hr rest

- Record $O_2$ SAT, $ETCO_2$ and end each trial
- Proceed with subsequent daily progressive increases of time on TM as follows
- Notify the pulmonary physician for proceeding with ventilator liberation if all criteria in the reassessment are met for spontaneous breathing trial.

**Tracheostomy Mask Progress:**

- 4 hrs BID (2 – 4 hrs rest interval in between)
- 8 hrs consecutive
- Up to 16 hrs consecutive, rest on ventilator support at night x 2 nights

When the patient can tolerate 16 hours x2 consecutive days with rest on ventilator support at night, proceed to 24 hours and obtain ABG after the 24-hour period. After obtaining the ABG, consult with pulmonary/critical care.

**Tracheostomy decannulation protocol**

- All patients undergoing decannulation will need to be evaluated by a Pulmonary or Critical Care Medicine consultant. The consultant will document the patient is ready to go ahead with the decannulation protocol.
- After evaluation, the Pulmonary consultant may proceed with decannulation off protocol with clear documentation to the reasons why.
- For patients unable to perform cough peak flow procedure, refer to the protocol designated for inadequate peak flow.
- Daily progress of the decannulation process will be documented by the Respiratory Care Practitioner (RCP) and conveyed to covering therapists during change of shift report.
- The RCP will maintain daily communication with the LIP and/or consultant of the progress of the decannulation process.
- Once the decannulation protocol has been completed, the RCP will notify the LIP of completion of the protocol, and the LIP will enter the decannulation order in Meditech. Please refer to the section on trach removal.

I. Criteria for Initiation of Protocol:

All patients must have all of the following: Afebrile; Hemodynamically stable; Clear or stable Chest X-ray; Controlled Secretions; Peak Cough Flow ≥160 L/m; Satisfactory on-going nutrition (low risk of aspiration); No clinical evidence of tracheal obstruction; Ability to tolerate a speaking valve.
Morning ABG immediately upon completion of the NOT study will be required.
The Respiratory Care Practitioner will determine that the criteria have been met. If the patient fails to meet the criteria, the LIP notified, and the protocol cancelled.

II. Respiratory Care Practitioners/Registered Nurses:

For ALL patients:

- Decrease tracheostomy cannula size to ≤ 6mm.
  - The first tracheostomy change (post initial surgical tracheostomy insertion), change will be done by an Otolaryngology, Pulmonary, or Critical Care Medicine physician.
- Nursing to monitor and record HR, BP, and temp Monitor VS q 4 hours for 24 hours with each tracheostomy change
- RT to monitor and record $O_2$ saturation, $ETCO_2$, and RR q4 hours for 24 hours with each trach change
- Observe trach site for bleeding
- RCP does trach change and documents in the Clinical Information System.
- RCP/RN will notify LIP if VS become unstable and/or $O_2$ Sat ≤ 92%

III. Criteria to begin plugging trials:

- VS stable (within 15% of baseline)
- For patients with no underlying lung disease: $O_2$ saturation ≥ 92% and $ETCO_2$ ≤ 45 mmHg
- For patients with underlying lung disease: Stable blood gases; With adequately compensated $PCO_2$ and pH; $O_2$ saturation ≥ 92% on room air or oxygen

IV. Decannulation

<u>Day 1</u>

- Plug tracheostomy
- Nursing to monitor and record HR, BP, and temp q 4 hours

- RT to monitor and record $O_2$ saturation, $ETCO_2$, and RR q 4 hours
- Plug up to 16 hrs, remove for sleep

Criteria for unplugging sooner than 16 hours if any of the following occur: Change in hemodynamics (BP 15% above or below baseline); Increased RR (15% above baseline); $O_2$ Sat < 92%; Increase in $ETCO_2$ of >20% over baseline; Increased HR (15% above baseline); Fever ≥ 100.8°F; Increase in pulmonary secretions; Stridor.
RCP documents plugging trial in the decannulation protocol documentation intervention and patient response/compliance in notes section of Clinical Information System.

Day 2
If successful with day 1 of 16 hour plugging, on day 2 plug trach and leave for 24 hours.

- Nursing to monitor and record HR, BP, and temperature q 4 hours
- RT to monitor and record $O_2$ saturation, $ETCO_2$, and RR q 4 hours
- Obtain overnight oximetry
- Obtain early am ABG according to criteria in section II

Day 3 (if required)

- If clinically necessary, may continue with tracheostomy plug ATC
- Nursing to monitor and record HR, BP, and temperature q 4 hours
- RT to monitor and record $O_2$ saturation, $ETCO_2$, and RR q 4 hours
- Obtain overnight oximetry (if not done previously)
- Obtain early am ABG according to criteria in section II (if not done previously)

Criteria for unplugging before 24 hrs: Any items listed in Section IV

RCP documents plugging trial and if appropriate that patient meets criteria for removal in notes section of the Clinical Information System.
If plugging trial fails RCP will notify LIP.
Any unsuccessful decannulation requiring replacement of the tracheostomy tube will require an Otolaryngology consult.
If a patient is moved to another unit as part of the decannulation process, RT staff will complete bedside hand-off of report.

Criteria for tracheostomy removal

- Decannulation will take place prior to 5pm unless approved by LIP
- Stable vital signs (within 15% of baseline)
- Oxygen saturation ≥ 92%
- $ETCO_2$ remains stable (within 5% of baseline)
- Overnight oximetry results **without** suggestion of sleep apnea.
- Overnight oximetry results **with** suggestion of sleep apnea:
    - RCP will leave trach in place and discuss further action with the LIP

If decannulation is decided, then the RCP will decannulate patient and place and securely tape sterile gauze over the patient's stoma.
RCP will then document procedure and patient response in the clinical information system.
Following decannulation, all patients will be monitored with pulse oximetry for the first 24 hours post decannulation, with vital signs monitored and recorded every 4 hours for 24 hours by nursing and RT (see above). If the patient is on telemetry during the decannulation process, he/she will be maintained on telemetry.

The LIP will be notified if patient experiences:

- Change in hemodynamics (BP 20% above or below baseline)
- Increased RR (15% above baseline)
- $O_2$ saturation < 92%
- Increase in $ETCO_2$ of >5% above baseline
- Increased HR (15% above baseline)
- Fever ≥ 100.8°F
- Increase in pulmonary secretions
- Development of stridor

If stable, patient may be discharged 48 hours after decannulation.
